# Features of α-HBDH in COVID-19 patients with different ages,outcomes and clinical types: a cohort study

**DOI:** 10.1101/2020.10.29.20222612

**Authors:** Haoming Zhu, Gaojing Qu, Hui Yu, Guoxin Huang, Lei Chen, Meiling Zhang, Shanshan Wan, Bin Pei

## Abstract

**Background:** Coronavirus disease-2019 (COVID-19) has spread all over the world and brought extremely huge losses. At present, there is no study to systematically analyse the features of hydroxybutyrate dehydrogenase (α-HBDH) in COVID-19 patients with different ages, clinical types and outcomes.

**Methods:** Electronic medical records including demographics, clinical manifestation, α-HBDH test results and outcomes of 131 hospitalized COVID-19 patients, with confirmed result of severe acute respiratory syndrome coronavirus 2 (SARS-CoV-2) viral infection, were extracted and analyzed.

**Results:** The α-HBDH value in ≥61 years old group, severe group and critical group, death group all increased at first and then decreased, while no obvious changes were observed in other groups. And there were significant differences of the α-HBDH value among different age groups (P<0.001), clinical type groups (P<0.001) and outcome groups (P<0.001). The optimal scale regression model showed that α-HBDH value (P<0.001) and age (P<0.001) were related to clinical type.

**Conclusions:** α-HBDH value increases in some COVID-19 patients, obviously in ≥61 years old, death and critical group, indicating that patients in these three groups suffer from more serious tissues and organs damage, higher α -HBDH value and risk of death. The obvious difference between death and survival group in early stage may provide a approach to judge the prognosis. The accuracy of the model to distinguish severe/critical type and other types is 85.84%, suggesting that α-HBDH could judge the clinical type of COVID-19 patients accurately. In brief, α-HBDH is an important indicator to judge the severity and prognosis of COVID-19.

## 1 Introduction

Since the outbreak of COVID-19 in December 2019, caused by severe acute respiratory syndrome coronavirus 2 (SARS-CoV-2), it has spread rapidly around the world. SARS-Cov-2 has attracted the attention of the global because of its high transmission ability, morbidity and mortality^**[1-4]**^. On January 30, the World Health Organization (WHO) identified COVID-19 as a public health emergency of international concern^**[5]**^. As of July 1, 2020, the number of confirmed cases of COVID-19 worldwide has reached 10357622, and the number of deaths has reached 508055^**[6]**^.

Lactate dehydrogenase (LDH) is one of the important enzymes in glycolysis and gluconeogenesis. It mainly catalyzes the transformation between lactic acid and pyruvate. Its enzymatic reaction is: pyruvate + NADH+H^+^⇌ lactic acid + NAD^+^. LDH consists of five isozymes composed of different combinations of H and M subunits: LDH1 (H4), LDH2 (H3M), LDH3 (H2M2), LDH4 (HM3) and LDH5 (M4). α-HBDH is tested by the α-ketoacid, a substrate, to determine the LDH activity. Additionally, the activity of LDH1 and LDH2 with more H subunits is described by α-HBDH activity because of the high affinity for this substrate to the H subunit in LDH. α-HBDH mainly distributed in the heart, brain, kidney and red blood cells, and the activity of the enzyme in the heart is more than half of the total enzyme activity. α-HBDH level increased in the progression of cor pulmonale, leukemia and tumor. Moreover, the extents of the increase and the tissue and organ injury were closely related, which can be used as an auxiliary diagnostic index^**[7-11]**^.

Compared with other pneumonia types, the α-HBDH level in COVID-19 patients was significantly higher, and the α-HBDH value of severe group was higher than that of non-severe group^**[12, 13]**^. When complicated with cardiovascular disease or gastrointestinal symptoms, the increase of α-HBDH in COVID-19 patients was much more significant as well^**[14, 15]**^. Cen Y et al. observed 1007 mild and moderate COVID-19 patients for 28 days. It was found that the higher the α-HBDH value, the greater the risk of progression to severe or critical type^**[16]**^. Zhang Gemin et al. divided 95 COVID-19 patients into four groups according to their α-HBDH level on admission. They found that the higher the α-HBDH value, the greater the proportion of severe cases, and the higher the risk of death or need for mechanical ventilation for patients, showing that the high α-HBDH level indicates an increased risk of further aggravation of the disease^**[17]**^. In this study, we analyzed the changes of α-HBDH values of COVID-19 patients with different ages, clinical types and outcomes. The effects of α-HBDH, age and gender on the clinical type of COVID-19 patients were quantified by the optimal scale regression model, so as to achieve the purpose of early judging the severity of the disease.

## 2 Methods

### 2.1 Study Population

Our study included individuals with complete examination data, 18 years old or above, who were hospitalized in Xiangyang No.1 People’s Hospital with a diagnosis of COVID-19. In the end, we included a total of 131 patients.

### 2.2 Study Design

This research project was a bidirectional observational cohort study. According to the diagnosis and treatment guidelines^[18]^, the patients were divided into mild group, moderate group, severe group and critical group. According to their age, the patients were divided into ≤40 years old group, 41-60 years old group and ≥61 years old group. According to the outcome, the patients were divided into death group and survival group. The distributions of α-HBDH median value were plotted with an 5-days interval (T1, T2, T3…Tn represented the time unit successively). The symptom onset data was designed as the first day of disease, the abnormal percentage, median and quartile interval of α-HBDH in different ages, outcomes and clinical types were calculated.

The study was approved by the ethics review board of Xiangyang No.1 People’s Hospital (No.2020GCP012) and registered at the Chinese Clinical Trial Registry as ChiCTR2000031088. Informed consent from patients has been exempted since this study is an observational cohort study that does not involve patients’ personal privacy.

### 2.3 Data Sources

Two groups (two researchers per group) extracted the data from hospital information system through a consistent data collection protocol and cross-checked. Gender, age, all α-HBDH test results, disease onset date, outcome, death date, etc. were collected. Two respiratory physicians classified the clinical types and then cross-checked the results. A third expert was involved when there was disagreement. The data were traced back to 23 January and followed up to 28 March, 2020.

### 2.4 Statistical Analysis

All statistical analyses were performed by SPSS 20.0. Continuous data in accordance with normality were represented by means and standard deviations, otherwise median (interquartile, IQR) was applied. Categorical data were described as frequency (%). The chi-square test was conducted to assess significance between groups. T-test was used to compare the quantitative data of normal distribution between the two groups, and the comparison of the quantitative data of non-normal distribution between the two groups was analyzed using Mann-Whitney U test. The correlation between two variables was tested by Spearman correlation test. The maximum α-HBDH value in the first 15 days, age and gender were regarded as independent variables and clinical types were regarded as dependent variables to build the optimal scale regression model.

## 3 Results

### 3.1 General Information

Up to February 28, 542 patients with suspected and confirmed COVID-19 have been admitted to our hospital. A total of 142 cases were positive for nucleic acid test, among which the data of 9 cases were incomplete, 2 cases were infants, thus 131 cases were included in this study finally, including 63 males and 68 females, aged 50.13±17.13 years old. Among them, there were 4 mild cases, 88 moderate cases, 18 severe cases and 21 critical cases. The average time from onset to admission, from onset to discharge, from onset to death, and length of hospitalization were 4.54±3.10, 26.87±9.19, 18.4±9.77 and 22.38±8.70 days respectively. In this study, 565 tests of α-HBDH were extracted from 37 laboratory indicators (including 24560 outpatient and inpatient examination results), accounting for 2.30% of the total test results.

### 3.2 α-HBDH in different age groups of COVID-19 patients

In ≤40 years old group, 41-60 years old group and ≥61 years old group, the α-HBDH median value was 123.17 (106.94-144.95) U/L, 150.49 (120.18-185.20) U/L and 221.59 (156.76-302.89) U/L, respectively, and the α-HBDH value abnormal percentage was 10.53%, 26.74% and 60.70%, respectively. The changes indicated that the α-HBDH median value in ≤40 years old group increased during T1-T2 and decreased after T2, and the normal range was T1 to T6; in 41-60 years old group, the α-HBDH median value decreased during T1-T6, and was in the normal range from T1 to T6; in ≥61 years old group, the α-HBDH median value increased during T1-T2 and decreased after T2, and the abnormal time interval was T2-T5 (Figure1, Table1).

**Table1.** α-HBDH in different ages of COVID-19 patients

| Period | $\leq 40$ years old(Group 1) | | | 41-60 years old(Group 2) | | | $\geq 61$ years old(Group 3) | | | P value |
| --- | --- | --- | --- | --- | --- | --- | --- | --- | --- | --- |
|  | Median<br>(IQR) | N | Abnormal<br>Percentage<br>(%) | Median<br>(IQR) | N | Abnormal<br>Percentage<br>(%) | Median<br>(IQR) | N | Abnormal<br>Percentage<br>(%) |  |
| First 30 Days | 123.17<br>(106.94-144.95) | 133 | 10.53% | 150.49<br>(120.18-185.20) | 172 | 33.33% | 221.59<br>(156.76-302.89) | 201 | 60.70% | P1<0.001,P2<0.001,<br>P3<0.001 |
| T1 | 131.88<br>(114.41-144.56) | 22 | 9.09% | 174.2<br>(133.20-193.85) | 21 | 35.00% | 175.65<br>(157.75-199.68) | 24 | 37.50% | P1=0.009,P2<0.001,<br>P3=0.306 |
| T2 | 144.25<br>(115.94-178.92) | 34 | 23.53% | 162.83<br>(138.83-193.96) | 40 | 32.26% | 258.66<br>(180.66-337.72) | 40 | 70.00% | P1=0.044,P2<0.001,<br>P3<0.001 |
| T3 | 114.44<br>(104.92-136.48) | 21 | 4.76% | 167.29<br>(128.72-211.43) | 31 | 17.24% | 233.23<br>(162.08-310.66) | 38 | 65.79% | P1<0.001,P2<0.001,<br>P3=0.002 |
| T4 | 120.07<br>(106.18-131.25) | 28 | 3.57% | 135.97<br>(118.59-156.72) | 29 | 25.93% | 232.36<br>(162.30-296.36) | 39 | 69.23% | P1=0.015,P2<0.001,<br>P3<0.001 |
| T5 | 113.17<br>(101.75-129.70) | 17 | 0.00% | 122.83<br>(111.43-178.23) | 27 | 12.50% | 200.05<br>(154.75-317.27) | 33 | 60.61% | P1=0.076,P2<0.001,<br>P3<0.001 |
| T6 | 111.57<br>(99.73-141.77) | 11 | 18.18% | 126.80<br>(111.96-160.15) | 24 | 26.74% | 172.36<br>(143.66-242.52) | 27 | 48.15% | P1=0.303,P2=0.003,<br>P3<0.001 |

P1: Group 1 vs Group 2, P2: Group 1 vs Group 3, P3: Group 2 vs Group 3, N: Total times of test in this period.

**Figure1.**
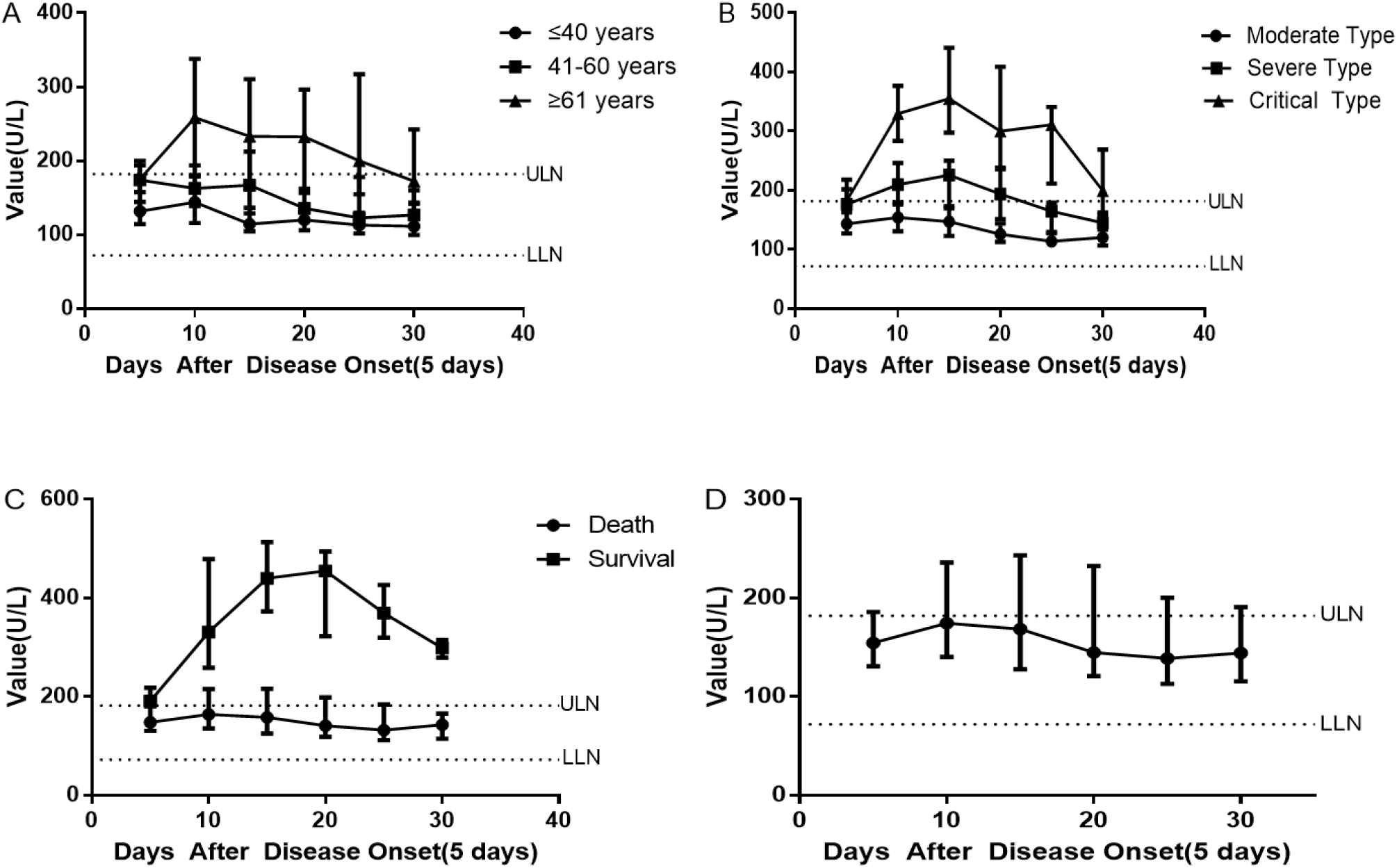
(A) the changes of α-HBDH in different age groups, (B) the changes of α-HBDH in different clinical type groups, (C) the changes of α-HBDH in different outcome groups, (D) the changes of α-HBDH in total course.

There were significant differences of the α-HBDH value in the first 30 days among the three age groups. Significant differences were observed in the α-HBDH value between ≤40 years old group and 41-60 years old group during T1-T4, between ≤40 years old group and ≥61 years old group during T1-T6, and between 41-60 years old group and ≥61 years old group during T2-T6. Differences were also significant in the α-HBDH value abnormal percentage among the three age groups (P<0.001). The age was correlated with α-HBDH value according to the Spearman correlation test (P<0.001), and the coefficient was 0.52.

### 3.3 α-HBDH in different outcome groups of COVID-19 patients

In survival group and death group, the α-HBDH median value was 147.80 (121.55-194.67) U/L and 337.18 (294.01-477.11) U/L, respectively, and the α-HBDH value abnormal percentage was 29.52% and 96.08%, respectively. The changes indicated that the α-HBDH median value in survival group increased during T1-T2, decreased after T2, and was in the normal range during T1-T6; while in death group, the α-HBDH median value increased during T1-T4 and decreased after T4, and the abnormal time interval was T1-T6 (Figure1, Table2).

**Table2.** α-HBDH in different outcomes of COVID-19 patients

| Period | Survival |  |  | Death |  |  | P value |
| --- | --- | --- | --- | --- | --- | --- | --- |
|  | Median(IQR) | N | Abnormal<br>Percentage<br>(%) | Median(IQR) | N | Abnormal<br>Percentage<br>(%) |  |
| First 30 Days | 147.80<br>(121.55-194.67) | 454 | 29.52 | 337.18<br>(294.01-477.11) | 51 | 96.08 | P<0.001 |
| T1 | 148.48<br>(130.85-180.97) | 61 | 24.59 | 191.11<br>(181.58-218.19) | 6 | 66.67 | P=0.014 |
| T2 | 165.02<br>(135.92-215.63) | 100 | 37.00 | 331.44<br>(258.94-479.36) | 13 | 100.00 | P<0.001 |
| T3 | 157.30<br>(125.68-214.05) | 81 | 33.33 | 440.35<br>(373.28-513.89) | 9 | 100.00 | P<0.001 |
| T4 | 141.14<br>(118.86-198.90) | 87 | 27.59 | 455.11<br>(322.65-426.79) | 9 | 100.00 | P<0.001 |
| T5 | 132.02<br>(112.06-184.22) | 69 | 27.54 | 369.98<br>(319.60-426.79) | 8 | 100.00 | P<0.001 |
| T6 | 142.90<br>(113.74-165.30) | 56 | 21.43 | 298.80<br>(279.30-315.31) | 6 | 100.00 | P<0.001 |

N: Total times of test in this period.

There were significant differences of the α-HBDH in the first 30days and every time unit during T1-T6. Differences were also significant in the α-HBDH value abnormal percentage among the two outcome groups (P<0.001). Spearman correlation test showed that there was a correlation between outcome groups and the α-HBDH value (P<0.001), and the correlation was 0.49.

### 3.4 α-HBDH in different clinical type groups of COVID-19 patients

In mild group, moderate group, severe group and critical group, the α-HBDH median value were 110.41 (105.67-120.25) U/L, 134.92 (114.40-163.23) U/L, 180.95 (144.02-231.01) U/L and 293.57 (209.53-368.64) U/L, respectively, and the abnormal percentage of α-HBDH value was 0.00%, 12.45%, 49.47% and 85.59%, respectively. The changes indicated that the α-HBDH median value of the moderate group increased during T1-T2 and decreased after T2, and was in the normal range during T1-T6; in the severe group, the α-HBDH median value increased during T1-T3 and decreased after T3, and the abnormal time interval was T2-T4; in the critical group, the α-HBDH median value increased during T1-T3 and decreased after T3, and the abnormal time interval was T1-T6 (Figure1, Table3). However, the mild group was excluded in statistical analysis for the reason that it only contained 4 cases.

**Table3.** α-HBDH in different clinical types of COVID-19 patients

| Period | Moderate Type(Group 1) |  |  | Severe Type(Group 2) |  |  | Critical Type(Group 3) |  |  | P value |
| --- | --- | --- | --- | --- | --- | --- | --- | --- | --- | --- |
|  | Median<br>(IQR) | N | Abnormal<br>Percentage<br>(%) | Median<br>(IQR) | N | Abnormal<br>Percentage<br>(%) | Median<br>(IQR) | N | Abnormal<br>Percentage<br>(%) |  |
| First 30 Days | 134.92<br>(114.40-163.23) | 273 | 12.45 | 180.95<br>(144.02-231.01) | 95 | 49.47 | 293.57<br>(209.53-368.64) | 118 | 85.59 | P1<0.001,P2<0.001,<br>P3<0.001 |
| T1 | 143.40<br>(127.33-178.20) | 41 | 19.51 | 176.28<br>(143.43-201.95) | 12 | 41.67 | 182.88<br>(163.28-218.19) | 10 | 50.00 | P1=0.046,P2=0.017,<br>P3=0.468 |
| T2 | 154.24<br>(130.94-179.86) | 64 | 20.31 | 211.30<br>(176.44-245.47) | 21 | 66.67 | 329.90<br>(281.99-396.87) | 23 | 100.00 | P1<0.001, P2<0.001,<br>P3<0.001 |
| T3 | 147.36<br>(123.09-169.97) | 49 | 14.29 | 222.56<br>(159.52-248.88) | 17 | 64.71 | 355.23<br>(297.57-440.91) | 18 | 100.00 | P1<0.001, P2<0.001,<br>P3<0.001 |
| T4 | 125.84<br>(113.13-143.89) | 54 | 3.70 | 194.13<br>(151.50-235.97) | 20 | 55.00 | 300.20<br>(238.58-408.87) | 20 | 100.00 | P1<0.001,P2<0.001,P<br>3<0.001 |
| T5 | 113.86<br>(108.60-129.70) | 37 | 10.81 | 160.69<br>(120.62-174.69) | 16 | 25.00 | 311.01<br>(211.61-340.84) | 23 | 82.61 | P1=0.008, P2<0.001,<br>P3<0.001 |
| T6 | 120.49<br>(106.79-143.89) | 28 | 0.00 | 143.82<br>(126.68-156.23) | 9 | 22.22 | 199.33<br>(158.84-268.88) | 24 | 66.67 | P1=0.306,,P2<0.001,<br>P3=0.007 |

P1: Group 1 vs Group 2, P2: Group 1 vs Group 3, P3: Group 2 vs Group 3, N: Total times of test in this period.

**Table4.**
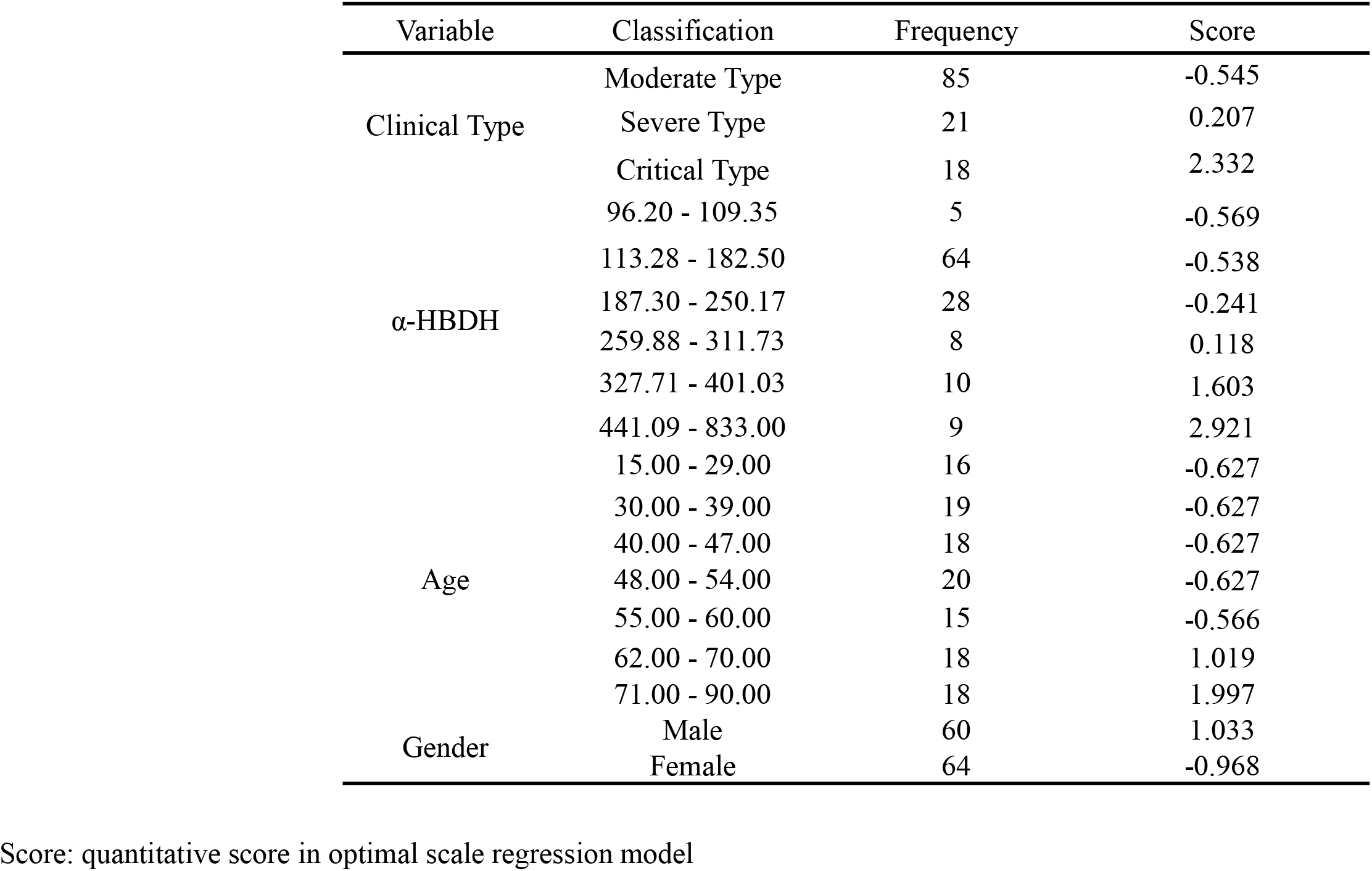
Variable Category and quantification score in optimal scale regression model

There were significant differences of the α-HBDH value in the first 30 days among the three clinical type groups. Significant differences were observed in the α-HBDH value between moderate type group and severe type group during T1-T6, between moderate type group and critical type group during T1-T6, and between severe type group and critical type group during T2-T6.

### 3.5 Optimal scale regression model based on this study

After excluded 4 mild cases, we built the optimal scale regression model based on the maximum α-HBDH value in the first 15 days, age, gender and clinical type. The adjusted R^2^ of the model was 0.659. The clinical type was significantly correlated with age (P<0.001) and α-HBDH value (P<0.001), but not with gender (P=0.337). The model expression was Q_levels = 0.648*Q_α-HBDH + 0.036*Q_gender + 0.271*Q_age (Q_levels, Q_ages, Q_α-HBDH and Q_gender represent the scale quantification scores of clinical type, age, α-HBDH value and gender, respectively). The comparison between the output type and the actual type quantification score was shown in the figure 2. According to the model, the output accuracy of moderate, severe and critical types was 81.45%. In order to discriminate moderate type and severe/critical type, the severe type and critical type were combined to the same category, and the accuracy was 85.84%.

**Figure2.**
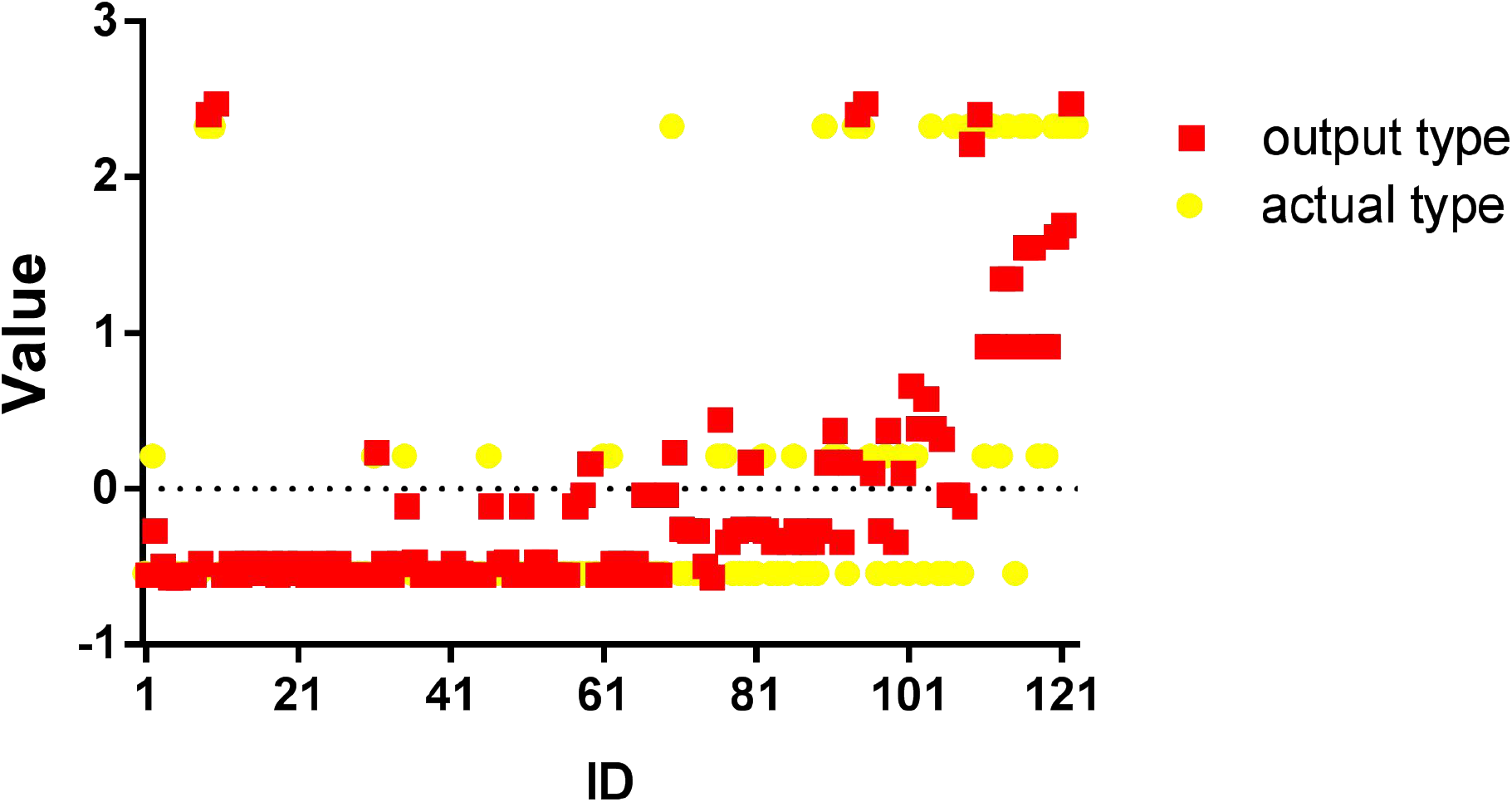
Comparison between the output type and the actual type quantification score in the model

## 4 Discussion

α-HBDH is one of the important enzymes in the process of glucose metabolism, which is widely distributed in various tissues and organs, especially in the heart, brain, kidney and red blood cells. COVID-19 mainly induces lung injury and causes damage to the heart and other tissues and organs, which results in the release of α-HBDH and the increase of α-HBDH in blood terminally^[17]^. The α-HBDH value in COVID-19 patients changed, it increased or significantly increased in some patients (Figure1). In different age groups, outcome groups and clinical type groups, there were significant differences in the distribution and abnormal percentage of α-HBDH. Age and outcome were significantly correlated with α-HBDH. Moreover, α-HBDH value may be related to the severity of COVID-19.

In ≥61 years old group and death group, the α-HBDH median value increased from T1, and reached a single peak in T2 (258.66 U/L) and T4 (455.11 U/L), respectively, which was significantly different from that in ≤40 years old group (peak value was 144.26 U/L), 41-60 years old group (peak value was 174.26 U/L) and survival group (peak value was 165.02 U/L). The abnormal interval of α-HBDH median value in ≥61 years old group and death group was T1-T5, T1-T6 respectively, which was significantly different from that in ≤40 years old group, 41-60 years old group and survival group, in which α-HBDH median value were all in the normal range. It shows that the older the age, the worse the outcome, the higher the α-HBDH value, and the longer the abnormal interval. The α-HBDH value in ≥61 years old group and death group was higher than that in other groups, indicating that the injury of heart, brain, kidney and other tissues and organs is more serious in elderly and death patients, which was consistent with previous study^[19]^. This may be related to the poor function of the immune system and being more sensitive to virus damage in elderly and seriously ill patients. This characteristic suggests that we should pay more attention to elderly patients and patients of which α-HBDH continues to increase significantly. According to the data distribution and statistical analysis, the α-HBDH value can distinguish the death and the survival. For example, during T1, patients with α-HBDH ≥180.97 U/L have a ≥25% chance of survival, and patients with α-HBDH ≥218.19 U/L have a ≥75% chance of death, which might have guiding significance for judgment of disease development in early stage. Enough cases and data could help us build a mathematic model and recognize the prognosis in advance better.

The α-HBDH median value of the mild group was in the normal range, without obvious change, indicating that the injury of tissue and organ injury in this type was slight. The same as mild group, α-HBDH median value in moderate group also distributed in the normal range. It increased firstly, and reached a peak (133.20 U/L) in T2, indicating that tissue and organ injury occurred immediately after the symptom onset in spite of the slight degree, and mainly occurred in the first 10 days. The changes of α-HBDH median value of severe group was similar to that of critical group, which increased during T1-T3 and decreased after T3. Their peak value was 222.56 U/L and 355.23 U/L appearing in T3, and recovered to normal range in T5 and T7, respectively. It shows that the more serious the illness, the higher the α-HBDH median value and peak value, the longer the abnormal interval, which may be related to the serious virus-induced acute lung injury, tissues and organs damage in severe and critical type groups. These characteristics are of great significance for us to judge the severity of the disease by α-HBDH.

The maximum α-HBDH value in the first 15 days, age and gender of all patients were selected to build the optimal scale regression model. α-HBDH <250.17 U/L and aged <60 years old was associated with moderate type, α-HBDH between 259.88-311.73 U/L and aged between 62-70 years old was associated with severe type, α-HBDH >327.71 U/L and aged >70 years old was associated with critical type. In this model, the output accuracy for clinical type was over 80%, which indicated that the model could distinguish the clinical classification based on our data well. α-HBDH and age could be used to discriminate the clinical type of COVID-19 patients if further verified by other data, which could help us to grasp the opportunity of treatment and reduce the risk of progression to severe and critical type in early stage.

However, this study has several limitations. All the patients in the study come from the single hospital, and the sample size is small. This study is a retrospective cohort study, which fails to detect and analyze the daily α-HBDH in patients and may lose some information.

## 5 Conclusion

α-HBDH value increases in some COVID-19 patients, obviously in ≥61 years old, death and critical group, indicating that patients in these three groups suffer from more serious tissues and organs damage, higher α-HBDH value and risk of death. The obvious difference between death and survival group in early stage may provide a approach to judge the prognosis. The accuracy of the model to distinguish severe/critical type and other types is 85.84%, suggesting that α-HBDH could judge the clinical type of COVID-19 patients accurately. In brief, α-HBDH is an important indicator to judge the severity and prognosis of COVID-19.

## Data Availability

Anyone who wishes to obtain the original data of this study with reasonable purposes can contact the correspondent author via email.

## References

1 Huang CL, Wang YM, Li XW, et al. Clinical features of patients infected with 2019 novel coronavirus in Wuhan, China. Lancet, 2020; 395 (10223): 497–506. doi:10.1016/S0140-6736(20)30183-5

2 Chen NS, Zhou M, Dong X, et al. Epidemiological and clinical characteristics of 99 cases of 2019 novel coronavirus pneumonia in Wuhan, China: a descriptive study. Lancet, 2020; 395 (10223): 507–13. doi:10.1016/S0140-6736(20)30211-7

3 Alexandra LP, Rebecca K, Lawrence OG. The Novel Coronavirus Originating in Wuhan, China: Challenges for Global Health Governance. JAMA, 2020. doi:10.1001/jama.2020.1097

4 Li Q, Guan XH, Wu P, et al. Early Transmission Dynamics in Wuhan, China, of Novel Coronavirus-Infected Pneumonia. The New England journal of medicine, 2020; 382 (13): 1199–1207. doi:10.1056/NEJMoa2001316

5 World Health Organization. Novel Coronavirus(2019-nCoV) Situation Report-11. Available at:https://www.who.int/docs/default-source/coronaviruse/situation-reports/20200131-sitrep-11-ncov.pdf?sfvrsn=de7c0f7_4

6 World Health Organization. Novel Coronavirus(2019-nCoV) Situation Report-163. Available at:https://www.who.int/docs/default-source/coronaviruse/situation-reports/20200701-covid-19-sitrep-163.pdf?sfvrsn=c202f05b_2

7 Patrick O, Laurent J, Olivier M, et al. Prognostic value of C-reactive protein and cardiac troponin I in primary percutaneous interventions for ST-elevation myocardial infarction. AM HEART J, 2006; 152 (6): 1161–1167. doi:10.1016/j.ahj.2006.07.016

8 Dissmann R, Linderer T, Schröder R. Estimation of enzymatic infarct size: direct comparison of the marker enzymes creatine kinase and alpha-hydroxybutyrate dehydrogenase. Am Heart J. 1998;135(1):1–9. doi:10.1016/s0002-8703(98)70335-7

9 Kemp M, Donovan J, Higham H, Hooper J. Biochemical markers of myocardial injury. Br J Anaesth. 2004;93(1):63–73. doi:10.1093/bja/aeh148

10 Gungor K, Philip L, Abdul BK. Serum alpha-hydroxybutyrate dehydrogenase levels in children with sickle cell disease. The American journal of pediatric hematology/oncology, 1981; 3 (2): 169–171. doi:10.1097/00043426-198100320-00010

11 Apostolov I, Minkov N, Koycheva M, et al. Acute changes of serum markers for tissue damage after ESWL of kidney stones. Int Urol Nephrol. 1991;23 (3):215–220. doi:10.1007/BF02550414

12 Zhao DH, Yao FF, Wang LL, et al. A Comparative Study on the Clinical Features of Coronavirus 2019 (COVID-19) Pneumonia With Other Pneumonias. Clinical infectious diseases, 2020; 71 (15): 756–761. doi:10.1093/cid/ciaa247

13 Dong YL, Zhou HF, Li MY, et al. A novel simple scoring model for predicting severity of patients with SARS-CoV-2 infection. TRANSBOUND EMERG DIS, 2020. doi:10.1111/tbed.13651

14 Li MY, Dong YL, Wang HJ, et al. Cardiovascular disease potentially contributes to the progression and poor prognosis of COVID-19.. Nutrition, metabolism, and cardiovascular diseases. NMCD 2020; 307(7): 1061–1067. doi:10.1016/j.numecd.2020.04.013

15 Zhang H, Liao YS, Gong J, et al. Clinical characteristics of coronavirus disease (COVID-19) patients with gastrointestinal symptoms: A report of 164 cases. Dig Liver Dis, 2020. doi:10.1016/j.dld.2020.04.034

16 Cen Y, Chen X, Shen Y, et al. Risk factors for disease progression in patients with mild to moderate coronavirus disease 2019-a multi-centre observational study. Clin. Microbiol. Infect, 2020; 26(9):1242–1247 doi:10.1016/j.cmi.2020.05.041

17 Zhang GM, Zhang J, Wang BW, et al. Analysis of clinical characteristics and laboratory findings of 95 cases of 2019 novel coronavirus pneumonia in Wuhan, China: a retrospective analysis. RESP RES, 2020; 21 (1): 74. doi:10.1186/s12931-020-01338-8

18 Nationan Health Commissiom of the People’s Republic of China. Diagnosis and treatment of corona virous disease-19 (8th trail edition). Available at:http://www.nhc.gov.cn/yzygj/s7653p/202008/0a7bdf12bd4b46e5bd28ca7f9a7f5e5a/files/a449a3e2e2c94d9a856d5faea2ff0f94.pdf

19 Zhang JJ, Dong X, Cao YY, et al. Clinical characteristics of 140 patients infected with SARS-CoV-2 in Wuhan, China. Allergy. 2020;75(7):1730–1741. doi:10.1111/all.14238

